# Inhibitory control and its modification in spider phobia – study protocol for an antisaccade training trial

**DOI:** 10.1101/2023.09.22.23295977

**Authors:** Anne Sophie Hildebrand, Fabian Breuer, Elisabeth Johanna Leehr, Johannes B. Finke, Leandra Bucher, Tim Klucken, Udo Dannlowski, Kati Roesmann

**Affiliations:** Department of Psychology, Clinical Psychology and Psychotherapy of the University of Siegen, 57072, Siegen, Germany; Institute for Translational Psychiatry of the University of Münster, 48149, Münster, Germany; Institute of Psychology of the University of Osnabrück, 49076 Osnabrück, Germany

**Keywords:** Antisaccade task, specific phobia, inhibitory control, skin conductance response, startle response

## Abstract

**Objectives:** Inhibitory control deficits are considered a key pathogenic factor in anxiety disorders. To assess inhibitory control, the antisaccade task is a well-established measure, assessing antisaccade performance via latencies and error rates. The present study follows three aims: (1) to investigate inhibitory control via antisaccade latencies and errors in an antisaccade task, and their associations with multiple measures of fear in patients with spider phobia (SP) versus healthy controls (HC), (2) to investigate the modifiability of antisaccade performance via a fear-specific antisaccade training in patients with SP and HC, and (3) to explore associations between putative changes in antisaccade performance in SPs and diverse measures of fear following the training.

**Methods:** Towards aim 1, we assess antisaccade latencies (primary outcome) and error rates (secondary outcome) in an emotional antisaccade task. Further, the baseline assessment includes assessments of psychophysiological, behavioral, and psychometric indices of fear in patients with SP and HCs. To address aim 2, we compare effects of a fear-specific antisaccade training with effects of a prosaccade training as a control condition. The primary and secondary outcomes are reassessed at a post-1-assessment in both SPs and HCs. Aim 3 employs a cross-over design and is piloted in patients with SP, only. Towards this aim, primary and secondary outcomes, as well as psychophysiological, behavioral, and psychometric measures of fear are reassessed at a post-2-assessment after the second training block.

**Conclusion:** This study aims to better understand inhibitory control processes and their modifiability in spider phobia. If successful, antisaccade training may assist in the treatment of specific phobia by directly targeting the putative underlying inhibitory control deficits. This study has been preregistered with ISRCTN (ID: <u>ISRCTN12918583</u>) on 28th February 2022.

## Background

Specific phobia constitutes the most prevalent anxiety disorder with a prevalence ranging from 1.5% to 14.4% in adults [1]. Specific phobia is characterized by excessive fear of a specific object (e.g. spiders) or a specific situation [2]. Real or expected confrontation with the fear-evoking stimulus elicits a direct fear reaction, thus prompting habitual avoidance of this stimulus. While specific phobia can be treated effectively [2], studies show that about one-third of patients do not benefit from common first-line treatments [3, 4]. A better understanding of the neurocognitive underpinnings of specific phobia as a model for anxiety disorders may pave the way for developing mechanism-informed targeted interventions that could add to the existing methods of treatment and address the problem of non-response to treatment.

Anxiety can be characterized by a fast initial shift in attention, in favor of threatening stimuli [5]. The Attentional-Control Theory distinguishes two systems that are involved in attentional control: a goal-driven top-down system and a stimulus-driven bottom-up system [6]. The presence of a fear-evoking stimulus is assumed to disrupt the balance of these systems in favor of bottom-up processing, causing a shift of attention towards the fear-evoking stimulus. From an evolutionary perspective, this is a highly adaptive mechanism preparing organisms for fast reactions towards (potential) threats [7]. Yet, if too sensitive, this mechanism may contribute to pathological anxiety, which is – amongst other aspects (e.g., avoidance, increased psychophysiological responses [8]) – characterized by an inability to adequately inhibit bottom-up-driven perceptual processes.

A well-established experimental task to assess inhibitory control is the antisaccade task [9]. In the antisaccade task, participants are instructed to look in the opposite direction of a peripherally presented visual stimulus. Indices of reduced inhibitory control are prolonged latencies of correct antisaccades and increased error rates (gaze at the visual stimulus, instead of the opposite direction). While the latency of correct antisaccades constitutes a measure of inhibitory control efficiency, the error rate reflects the effectiveness of inhibitory control performance [10]. Antisaccades have been used in a variety of different psychiatric populations to assess inhibitory control. Perhaps most prominently, the antisaccade task has been applied in the research of psychotic and schizophrenic disorders [9], however it has also been used to study inhibitory control in obsessive-compulsive disorder [11], post-traumatic stress disorder [12], binge-eating disorder [13], attentional-deficit-hyperactivity disorder [14], and alcohol use disorder [15]. Additionally, research has identified reduced inhibitory control efficiency, indexed by prolonged latencies of correct antisaccades, in a multitude of highly anxious, yet subclinical participants [16, 17]. Reduced inhibitory control at high levels of (dispositional) anxiety is further reported in a meta-analysis investigating the association between inhibitory control and anxiety in a multitude of different tasks [18]. In line with this, it is considered a risk and maintaining factor of pathological anxiety [19]. Surprisingly, there are no studies investigating inhibitory control via antisaccade performance in patients with specific phobia. Thus, the profile of probable inhibitory control deficits in patients with specific phobia and its association with other indices of fear, like avoidance behavior and psychophysiological responses, remains elusive. Furthermore, it is unclear, whether patients with specific phobia show impaired inhibitory control only in response to phobia-relevant stimuli or if they exhibit a general deficit in inhibitory control.

In previous studies, antisaccade training improved antisaccade performance in non-clinical and clinical samples [20, 21]. In a sample of patients with binge eating disorder, antisaccade training improved not only antisaccade performance in response to disorder specific stimuli (i.e., high-caloric food), but was also associated with a significant decrease of binge eating incidents. However, no changes could be found in psychometric measures associated with binge eating (e.g., food craving [21]). To date, there are no studies investigating the trainability of antisaccade performance in specific phobia.

To further elucidate the role of inhibitory control performance in specific phobia, the present study investigates antisaccade performance in patients with spider phobia (SP) compared to healthy controls (HC). As the literature on (dispositional) anxiety and inhibitory control mainly indicates inhibitory control deficits with regard to efficiency rather than effectiveness, antisaccade latency serves as the primary outcome. Antisaccade error rate (as a marker of inhibitory control effectiveness) is investigated as a secondary outcome. Our study follows three main aims: First, patients with spider phobia (as a model anxiety disorder) will be investigated via the antisaccade task regarding potential inhibitory control deficits and their associations to stimulus category (phobia-related versus neutral), psychophysiological (heart rate, skin conductance, startle-response), behavioral and psychometric measures of fear (aim 1). This aim employs a proof-of-concept mixed factorial design with the between subject factors group (SPs vs. HCs) and the within-subject factor stimulus material (phobia-related vs. neutral). Second, the modifiability of inhibitory control through a fear-specific antisaccade training will be piloted (aim 2). This aim employs an exploratory randomized controlled parallel group design with two training conditions (antisaccade training vs. control) that will be realized in SPs and HCs. Changes in antisaccade performance from baseline to post-1-assessment, and their dependence on stimulus material will assessed. Third, putative changes in inhibitory control performance following the training will be investigated regarding associations to changes in psychophysiological, behavioral, and psychometric measures of fear in SPs, therefore exploring possible therapeutic utility (aim 3). This aim employs an exploratory cross-over design in SPs only, i.e. patients will switch training conditions after the post-1-assessment, and will be assessed regarding antisaccade performance, psychophysiological (heart rate, skin conductance, startle-response), behavioral and psychometric measures of fear at post-2-assessment.

## Methods

### Participants

Participants with a specific phobia of the animal subtype (spiders) and HCs are recruited at the University of Siegen and from the general population. Interested participants are invited to a short telephone screening (∼ 10 minutes) and then enrolled in a more extensive clinical interview (∼ 1 hour) via telephone to assess further eligibility, conducted by trained research assistants. SPs have to fulfill the criteria of specific phobia according to the Diagnostic and Statistical Manual of Mental Disorders-IV (SCID-I, Section F [22], while HCs must not fulfill the criteria of spider phobia and not exceed a value of 19 points in the Spider Phobia Questionnaire (SPQ) [23]. Inclusion criteria for both groups are an age between 18 and 65 years, normal or corrected to normal vision, as well as normal hearing. Exclusion criteria are any psychiatric condition (except spider phobia, as well as a mild to moderate depressive episode in the past SPs) assessed via the Mini-International Neuropsychiatric Interview (MINI) [24]. Furthermore, any history of neurological disorders, current intake of Benzodiazepines or Barbiturates, astigmatism, hearing disorders, subjective auditory hypersensitivity, regular nicotine consumption (> 5 cigarettes/day) or known allergy to bites of insects and arachnids lead to exclusion.

### Sample size

Based on two studies using the antisaccade task in clinical samples, we calculated two a-priori power analyses using G*Power 3.1 [25] for ANOVAs (Analyses of Variance) to detect large effect sizes (Coheńs *f* = .40, *α* = .05, power = .85) in our primary outcome (i.e. antisaccade latencies) at baseline (aim 1) [12] and for the training effect (aim 2) [21]. Results indicated a required total sample size of 59 participants (30 per group).

### Procedure

#### Overall procedure

After an initial telephone screening, eligible participants are invited to provide written informed consent and participate in a clinical interview via telephone. If they fulfill eligibility criteria, they are invited to a laboratory appointment at the University of Siegen and asked to fill in an online battery of questionnaires, including the SPQ [23], beforehand via LimeSurvey 3.28.18 [26]. A schedule of enrollment, intervention and assessment can be found in Figure 1.

**Figure 1.**
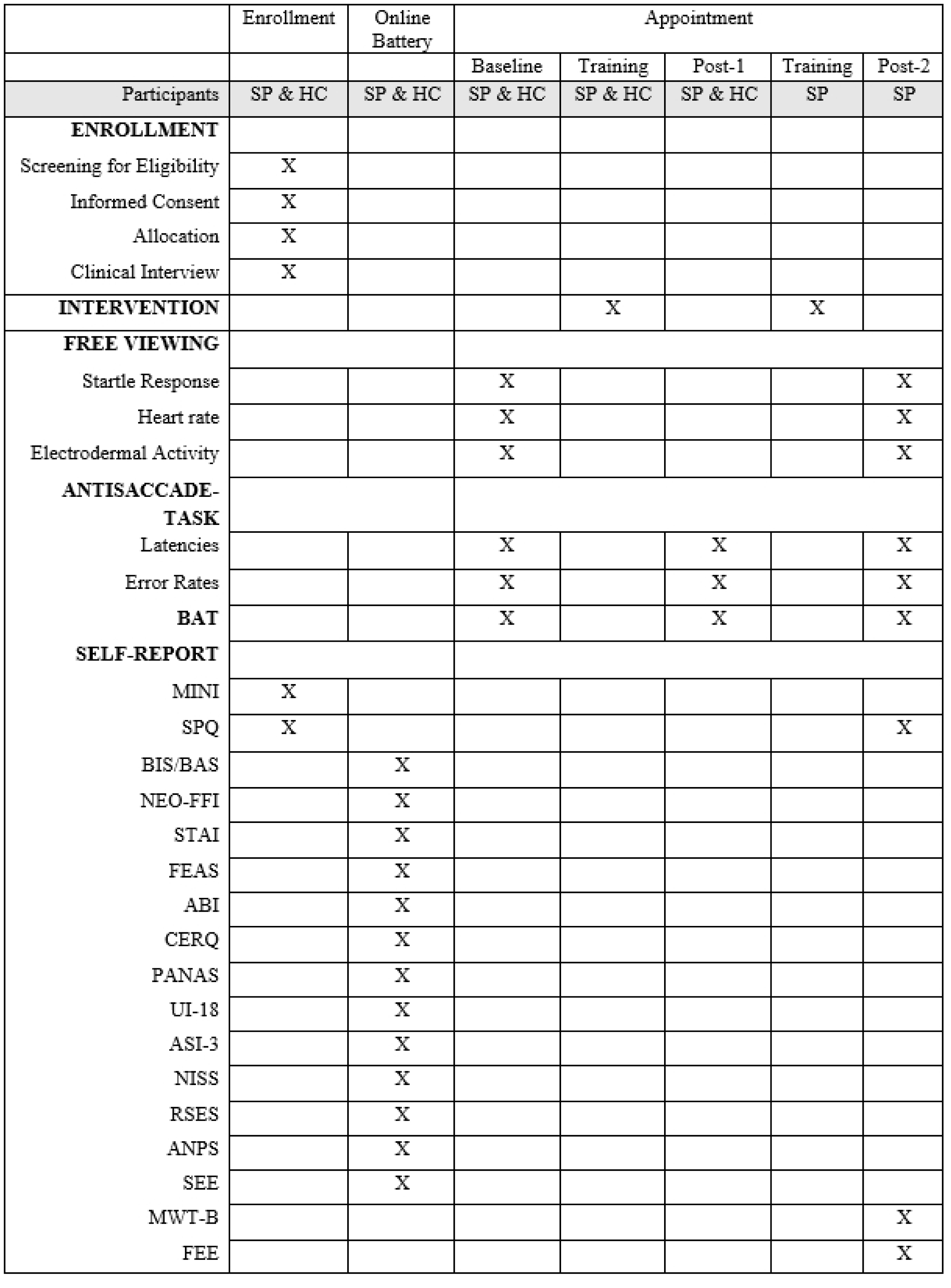
Schedule of Enrollment, Intervention, and Assessment.

##### Note

Assessment schedule, including all outcome measures. An X in the corresponding box indicates that assessment takes place at a certain time point. BAT: Behavioral Avoidance Test. MINI: Mini International Neuropsychiatric Interview [24]. SPQ: Spider Phobia Questionnaire [23]. BIS: Behavioral Inhibition System [27]. BAS: Behavioral Approach System [27]. NEO-FFI: NEO Five-Factor Inventory [28]. STAI: State-Trait-Anxiety-Inventory [29]. FEAS: Questionnaire on Disgust and Fear of Spiders [30]. ABI: inventory of coping with anxiety [31]. CERQ: Cognitive Emotion Regulation Questionnaire [32]. PANAS: Positive and Negative Affect Schedule [33]. UI-18: Intolerance of Uncertainty [34]. ASI-3: Anxiety Sensitivity Index [35]. NISS: Need Inventory of Sensation Seeking [36]. RSES: Rosenberg Self-Esteem Scale [37]. ANPS: Affective Neuroscience Personality Scales [38]. SEE: Scales for experiencing emotions [39]. MWT-B: Multiple-Choice Vocabulary Intelligence Test [40]. FEE: Disgust Responsiveness [41].

The laboratory appointment begins with a baseline assessment, including a behavioral avoidance test (BAT), a free-viewing task, and the antisaccade task (all of which are described below). This part of the study corresponds to aim 1 (see *background* and Figure 2). Then, participants will complete the antisaccade training or the control condition (prosaccade training), according to a randomization scheme. After training, the antisaccade task and the BAT will be repeated (post-1-assessment), and the study ends for the HCs. This part of the study corresponds to aim 2 (see *background* and Figure 2). The SPs will then complete a second training. Here, participants switch training conditions. After the second training, the antisaccade task, the free-viewing task, the BAT, and the SPQ will be repeated (post-2-assessment). This part of the study corresponds to aim 3 (see *background* and Figure 2).

**Figure 2.**
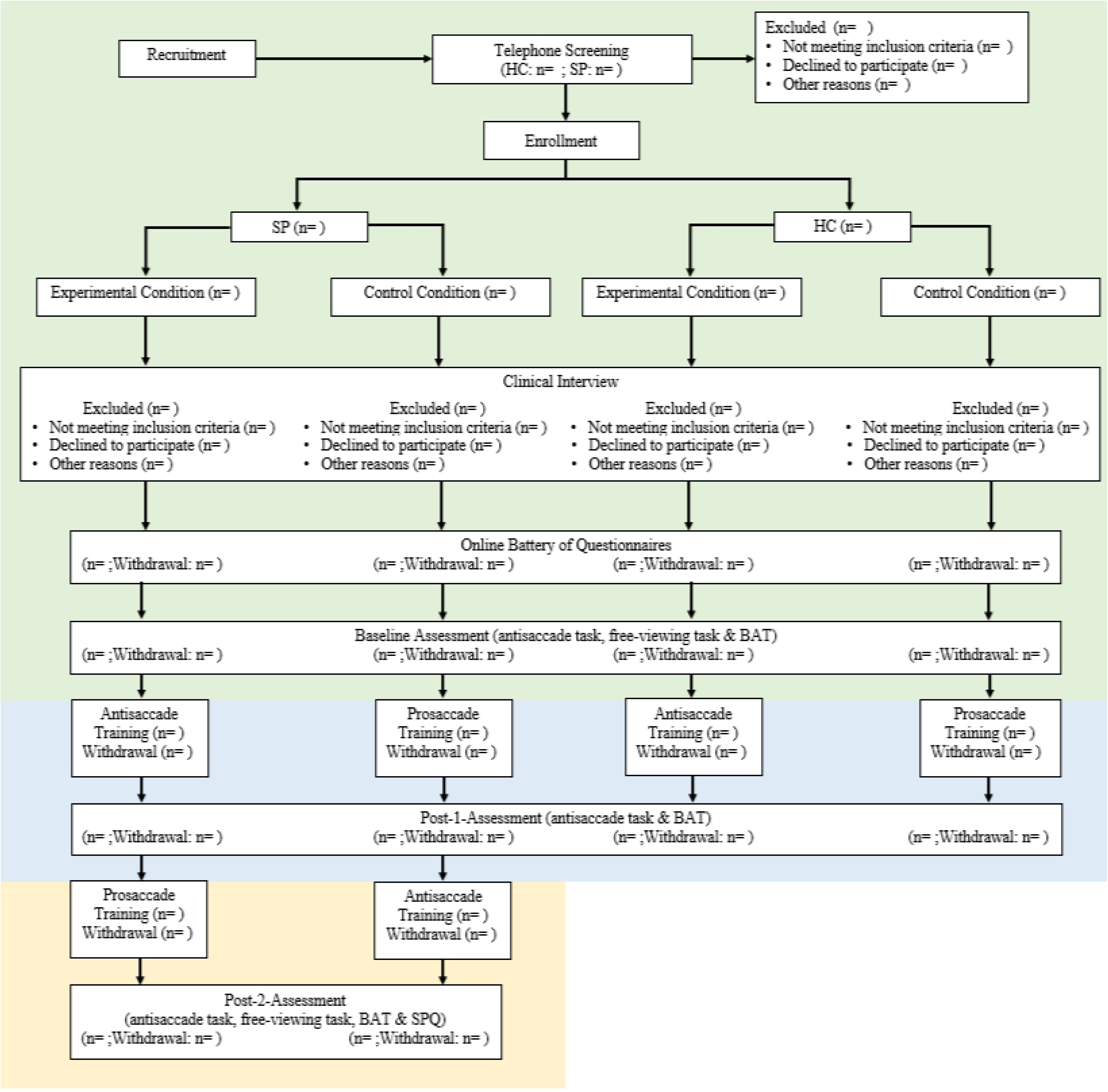
The Consort Flow Diagram.

##### Note

BAT: Behavioral avoidance test. HC: Healthy control participants. n: Number of participants. SP: Participants with spider phobia. SPQ: Spider phobia questionnaire. Key assessments to address aim 1 are shaded in green (baseline-assessment). Additionally, the intervention (antisaccade and prosaccade training) and the post-1-assessment are relevant for aim 2 (i.e., areas shaded in green and blue). In addition to assessments targeting aim 1 and 2, a second intervention and the post-2-assessments is relevant for aim 3 (i.e., areas shaded in green, blue and yellow). Note: Aim 3 refers to participants with spider phobia, only.

#### Laboratory assessments

##### Antisaccade Task

In the antisaccade task, ten stimuli, created and used by Kolassa and colleagues in previous studies [42], are presented. Stimulus material comprises five phobia-related pictures (black schematic pictures of spiders) and five neutral pictures (black schematic pictures of flowers), as depicted in Figure 3, to assess the stimulus-dependency of predicted differences in inhibitory control functions.

**Figure 3.**
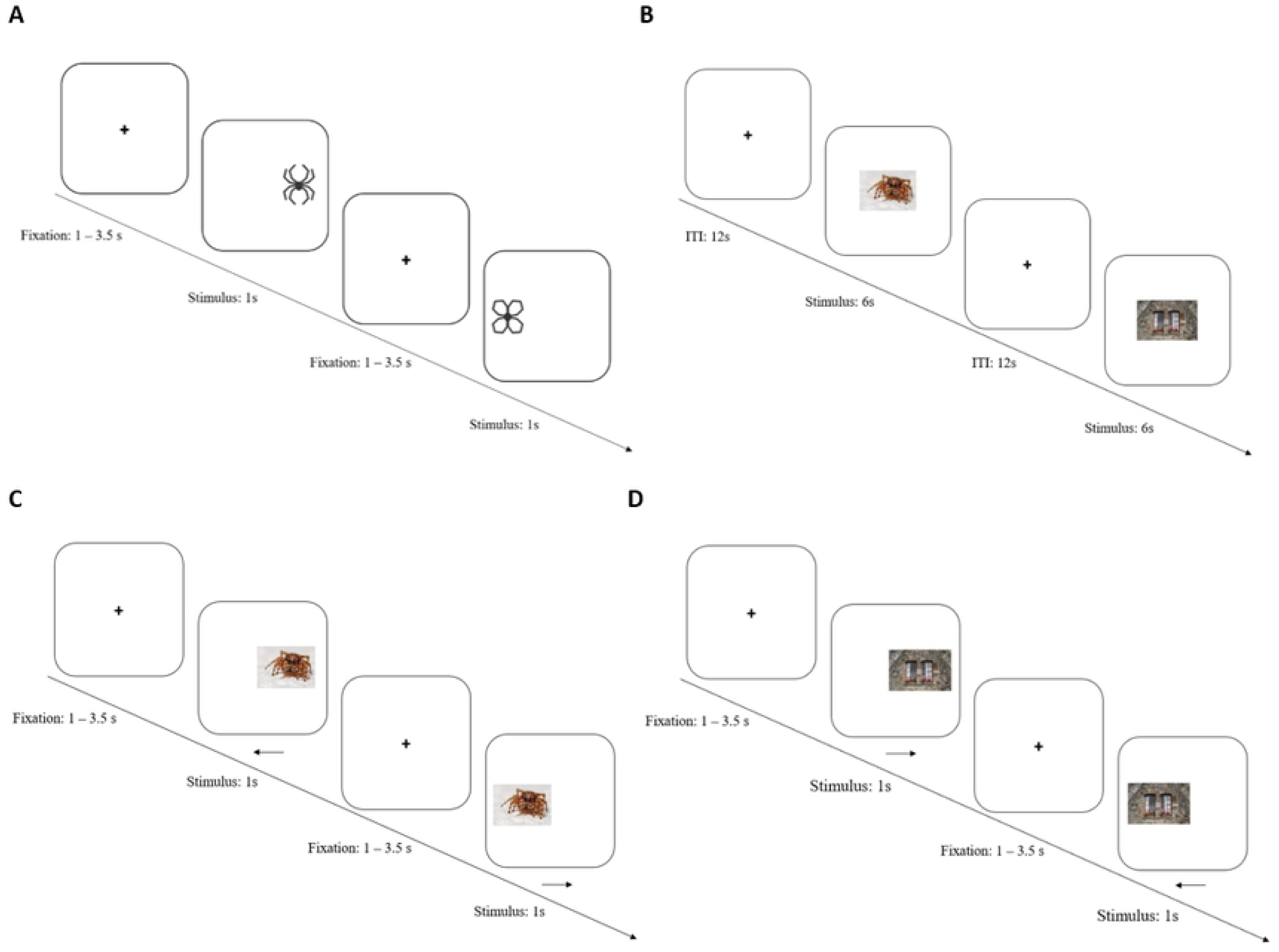
Overview of Tasks and Intervention.

###### Note

A: Antisaccade task, employs stimuli from [42]. B: Free-viewing task, uses phobia-related stimuli from GAPED [43]. C: Antisaccade training, uses additional phobia-related stimuli from GAPED. Arrows show the instructed viewing direction of the participants. D: Prosaccade training, uses neutral stimuli from GAPED. Arrows show the instructed viewing direction of the participants. ITI: Inter-Trial-Interval. s: Seconds.

The antisaccade task is conducted at baseline, post-1- and post-2-assessment. For each assessment, participants are placed in a darkened room. Eye movements are recorded via Eye-Link 1000 plus (SR Research Ltd., Canada) at a sampling rate of 1000 Hz. For each trial, a black fixation dot (0.25° x 0.25°) is presented on a white screen, with 84 cm distance to the participant, for a period of 1.5 - 3.5 seconds. After the fixation period, the dot vanishes, and the visual stimulus is immediately presented (so-called step-paradigm). The visual stimulus (3° x 2.25°) is randomly presented at 12° to the left or right of the visual field for one second.

Phobia-related and neutral stimuli are presented equally often in a randomized order. At baseline, the task includes five blocks as proposed by Antoniades and colleagues [44]: three antisaccade blocks (each including 40 antisaccades, i.e. 20 antisaccades to phobia-related and 20 antisaccades to neutral stimuli, each stimuli will be shown 4 times), as well as two prosaccade blocks (each including 60 prosaccades, i.e. 30 prosaccades to phobia-related and 30 prosaccades to neutral stimuli, each stimuli will be shown 6 times), one preceding and one following the antisaccade blocks. Antisaccade tasks at post-1- and post-2-assessment include only three blocks of antisaccades (each including 40 antisaccades), as prosaccades are already assessed at baseline, expenses for participants should be reduced and outcomes are only related to antisaccade performance. Each block is followed by a one-minute break.

Assessment of antisaccade performance will be relevant for aims 1 – 3 of the study.

##### Free-Viewing Task and Valence and Arousal Rating

In the free-viewing task, 24 pictures of the *Geneva Affective Picture Database (GAPED)* [43] are presented. The selected stimulus material comprises eight neutral pictures (inanimate objects), eight generally aversive but phobia-unrelated pictures (animal mistreatment), and eight phobia-related pictures (naturalistic spiders), respectively. Pictures are presented in two blocks of trials, each block depicting all of the 24 pictures in a randomized order. Via ratings provided by the authors of the database [43], negative pictures and pictures of spiders were matched regarding valence (negative: *M* = 21.35, *SD* = 8.78; spider: *M* = 20.46, *SD* = 7.77, *t*(15) = 1.00, *p* = .83) and arousal (negative: *M* = 66.30, *SD* = 5.18; spider: *M* = 66.99, *SD* = 6.41, *t*(15) = 1.02, *p* = .84), while neutral pictures had higher valence (*M* = 50.54, *SD* = 1.65) and lower arousal ratings (*M* = 21.77, *SD* = 6.01). Pictures are presented for six seconds each, followed by a twelve-second inter-trial-interval, as depicted in Figure 3.

While the pictures are presented, psychophysiological measures are assessed. To assess participants’ startle response, an acoustic stimulus (white noise, 50 ms, 105 dB(A), instantaneous rise time) is presented via headphones. This stimulus is presented 18 times during visual stimulus presentation for each block, that is six times per visual stimulus category, in a randomized order. The startle-eliciting stimulus occurs randomly between 4 and 5.5 seconds after visual stimulus onset. During inter-trial intervals, startle responses are elicited 6 times per block in a randomized order between 7.5 and 9.5 seconds after inter-trial-interval onset. Before each block, the startle-eliciting stimulus is presented 6 (first block) or 3 times (second block), respectively, to ensure initial habituation. Startle magnitude is measured by activity of the musculus orbicularis oculi (unilateral) via electromyography (EMG), utilizing a Biopac MP160 system (Biopac Systems, Inc., USA) with H124SG electrodes (Kendall). Design of startle assessment closely follows established methods [45]. Heart rate will be derived from continuously recorded electrocardiogram (ECG) utilizing a Biopac MP160 system with EL503 electrodes (Biopac). To assess skin conductance responses, electrodermal activity [46, 47] is measured utilizing a Biopac MP160 system with EL507 electrodes (Biopac) at the thenar/hypothenar of the non-dominant hand.

Psychophysiological measures are assessed in the free-viewing task at baseline and post-2-assessment and will be relevant to address aims 1 and 3 of the study.

Directly after the free-viewing task, each picture is presented again, and participants are asked to rate each picture in terms of valence and arousal on a visual analog scale. Pictures and the sequence of the valence and arousal ratings are randomized between participants. Ratings are used to describe stimulus-characteristics based on the study sample.

##### Behavioral Avoidance Test (BAT)

Avoidance behavior is assessed by an in vivo BAT. Behavioral avoidance is indexed by the final distance (in cm) between the participant and a spider (brachypelma auratum). For a detailed description, also see [48]. BAT is conducted at baseline, post-1- and post-2-assessment and will be relevant to address aims 1 and 3 of the study.

#### Intervention

##### Experimental Condition (Antisaccade Training)

The intervention in this trial will constitute an antisaccade training. Participants must look at the center of a screen. As depicted in Figure 3, stimuli will be presented for one second left or right in the periphery field of vision with a visual angle of 12°, after a fixation period of 1 - 3.5 seconds, based on recommendations by Antoniades and colleagues [44]. Participants are instructed to look at the mirrored position on the screen. Stimuli will constitute ten (additional) spider stimuli from the *GAPED* [43], which are shown in random order. We opted for phobia-related stimuli because the training of inhibitory control would be expected to be maximal in this condition. The training includes two blocks with 80 antisaccades each, separated by a pause of one minute. Adherence to interventions will be improved by carefully instructing participants before each training block. Implementation of the described training (including the control condition described below) will be relevant to address aims 2 and 3 of the study.

##### Control Condition (Prosaccade Training)

To examine the effect of antisaccade training on antisaccade performance, we will compare the effects of the antisaccade training with the effects of a prosaccade training on antisaccade performance. The prosaccade training was employed to control for potential fatigue effects caused by voluntary eye movements in general rather than antisaccades specifically.

In the control condition, as depicted in Figure 3, participants are instructed to look at stimuli that appear left or right in the periphery field of vision for one second after a fixation period of 1 - 3.5 seconds. Ten neutral stimuli from the *GAPED* [43] are used. In the control condition, we employ neutral stimuli rather than the phobia-related stimuli employed in the antisaccade training to prevent potentially beneficial effects of perceptual exposure on antisaccade performance. The training includes two blocks with 80 prosaccades each, separated by a pause of 60 seconds.

### Outcome

The primary outcome of this study is correct antisaccade latency, measured at baseline, post-1- and post-2-assessment in the antisaccade task. The secondary outcome is antisaccade error rate, also measured at baseline, post-1- and post-2-assessment in an antisaccade task.

Psychophysiological measures include startle response, heart rate, and skin conductance level, measured at baseline and post-2-assessment in a free-viewing task. As a behavioral assessment, the final distance in the behavioral avoidance test (BAT) will be conducted at baseline, post-1- and post-2-assessment. For psychometric assessment, the SPQ will be conducted at baseline and post-2-assessment.

Exploratory analyses will be conducted to pilot a fear-specific antisaccade training. Therefore, potential changes in antisaccade performance will be associated with changes in psychophysiological, behavioral, and psychometric measures of fear.

To characterize the sample, we will additionally obtain a range of self-report measures, described in Figure 1. All self-assessment questionnaires will be given out in the German version.

### Analysis

#### Preprocessing

##### Eye Tracking

To detect saccadic eye movements, conservative thresholds are used. Saccades are defined as eye movements with a velocity > 35°/sec, an acceleration > 9500°/sec^2^ and an amplitude > 3°. For the trial to be classified as valid, onset latency of the first saccadic eye movement after stimulus onset must be greater than 70 ms. Starting coordinates of the eye movement must fall within a square of 5° x 5° around the fixation stimulus. Additionally, blinks must be absent. Trials with no recorded eye movements after stimulus onset are also classified as invalid. A correct antisaccade (secondary outcome) is defined as a valid saccade in the opposite direction of the peripheral visual stimulus. Antisaccade latency (primary outcome) is defined as the time difference between stimulus onset and initiation of the correct antisaccade. Participants will be excluded from analysis if > 50% of maximal possible trials are invalid.

##### Psychophysiological data

ECG data is pre-processed using the PhysioData Toolbox (Version 0.6.3; [49]. The signal is filtered using a bandpass filter with a low cut-off at 1 Hz and a high cut-off at 50 Hz. From the ECG signal, R-peaks are retrieved, which serve to calculate heart rate measures. The ECG signal will be visually inspected for artifacts, as well as erroneous or missed R-peaks. Missed R-peaks will be added manually, erroneous R-peaks will be deleted. Trials with artifacts in the ECG signal will be discarded.

Skin conductance data is pre-processed using the PhysioData Toolbox. The raw signal is filtered and resampled to a 20 Hz time-vector spanning the length of the filtered signal, using spline interpolation. By first applying a moving minimum filter, followed by smoothing the result applying a Gaussian kernel, the tonic component is calculated. The phasic component is calculated by applying a 1st order high pass Butterworth filter of 0.5 Hz. The signal will be visually inspected for artifacts. Trials with artifacts in the SCL signal will be discarded.

The raw EMG signal is filtered using a band-pass filter ranging from 10 to 500 Hz, as well as rectified and smoothed, using a low-pass resistor-capacitor filter, utilizing a time constant of 10 ms. A C++-based software tool will be used to identify peak responses in the rectified and integrated EMG signal. Additionally, the signal will be visually inspected for artifacts (e.g., preceding blinks immediately before the startle probe), as well as erroneously detected response peaks. Trials with artifacts in the EMG signal will be discarded, erroneously detected response peaks will be corrected manually. Startle response amplitude is defined as the difference between EMG peak and baseline amplitude in a 200 ms response window.

#### Statistical Analyses

In the following, planned analyses will be described in brief. Further details on statistical analyses for <u>aim 1</u> and <u>aim 2</u>, can be found in the detailed statistical analyses protocols for each aim. For further details on aim 3, a statistical analyses protocols will be preregistered with Current Controlled Trials (ID: <u>ISRCTN12918583</u>).

Corresponding to aim 1 of the study, baseline differences in antisaccade performance between participants with SP and HCs will be investigated using general linear modeling with antisaccade latency and antisaccade error as outcome respectively. Diagnosis will be entered as a between-subject-factor, stimulus category as a within-subject factor, resulting in a 2×2 design. To investigate associations between multimodal measures of fear (psychophysiological responses, BAT and SPQ scores) and antisaccade performance in SPs, multiple regression analyses will be conducted with the primary and secondary outcome as dependent variables, respectively. If relevant demographic or psychometric measures differ between groups, these measures will be entered as covariates (for <u>statistical analysis plan</u> see ISRCTN12918583).

Corresponding to aim 2 of the study, general linear modelling will be conducted with diagnosis and intervention (antisaccade training vs. prosaccade training) as between-subject factors, as well as time and stimulus category as within-subject factors, resulting in a 2×2×2 design, including antisaccade latency and error rate as outcomes. Again, potential confounds showing group differences will be entered as covariates (for <u>statistical analysis plan</u> see ISRCTN12918583).

Corresponding to aim 3 of the study, exploratory analyses will be conducted in SPs, associating changes in antisaccade performance (antisaccade latency and error rate, pre vs. post-2) with changes in physiological responses to fear-specific stimuli, avoidance behavior, and self-reported fear of spiders (baseline vs. post-2). Additionally, antisaccade latencies and error-rates will be investigated via general linear modelling with intervention (antisaccade training first vs. prosaccade training first) as between-subject factors, as well as time (baseline, post-1, post-2) and stimulus category as within-subject factors, resulting in a 2×2×3 design. BAT scores will be investigated via general linear modelling with intervention (antisaccade training first vs. prosaccade training first) as between-subject factors, and time (baseline, post-1, post-2) as within-subject factor.

For all analyses significance levels will be set to *p* = .05.

## Discussion

The present study is designed to compare the antisaccade performance of SPs to a HC group and to examine the putative potential of an antisaccade training with phobia-related stimuli to increase inhibitory control in specific phobia. It also aims to explore associations between antisaccade performance and multimodal psychophysiological, behavioral, and psychometric measures of fear.

Focusing on the comparison of SPs and HCs (aim 1), a decreased antisaccade performance in SPs would yield evidence for reduced inhibitory control functions in SPs. This finding would extend previous findings, which have yielded evidence for associations between antisaccade performance and subclinical anxiety levels [16, 17], to clinically anxious individuals. Thereby, such findings would provide further evidence for a key role of inhibitory control processes in the pathogenesis and maintenance of specific phobia. Such evidence from the study of antisaccade performance would complement previous evidence from the field of fear conditioning, which has suggested deficient inhibitory learning processes and reduced inhibition of fear responses to safe stimuli in patients with pathological anxiety [50] and non-responders to exposure therapy in spider phobia [51]. As spider phobia is used as a model disorder for anxiety disorders more broadly, results could also serve as foundation for research on a wider range of anxiety disorders.

To estimate the clinical relevance of putative deficits in antisaccade performance (i.e., inhibitory control), it seems pivotal to shed light to its link with symptoms of fear. We therefore investigate relationships between antisaccade performance and multimodal measures of fear (e.g., psychophysiological fear responses, avoidance behavior as assessed via the BAT, and self-reported symptoms as assessed via the SPQ). The finding of significant associations could broaden our knowledge on the mechanistic basis of specific aspects of the fear response and form the basis for future research on a potential (causal) role of inhibitory control on fear-associated symptoms.

If a lower inhibitory control is found in patients with pathological anxiety, an antisaccade training could increase antisaccade performance as seen in previous studies investigating patients with other disorders [20, 21]. Aim 2 of this study targets this hypothesis. A positive result would suggest that an antisaccade training does increase inhibitory processes, which may in turn be a factor related to treatment outcomes in specific phobia (for review, see [52]. If this is the case, the generalizability of improved antisaccade performance to clinically relevant measures of fear and anxiety would be of importance. According to aim 3 of this study, we therefore investigate whether changes in antisaccade performance generalize to other measures of fear. Positive results regarding aim 2 and potentially also aim 3 would point towards the potential of the antisaccade training as a mechanism-informed targeted intervention for anxiety disorders [53]. As such, it might yield add-on effects to existing treatments and increase treatment response.

To conclude, findings of this study on patients with spider phobia (as a model for anxiety disorders more broadly) can strengthen our understanding of the mechanistic underpinnings of anxiety disorders and potentially pave the way for new targeted interventions.

## Data Availability

No datasets were generated or analysed during the current study. All relevant data from this study will be made available uponrequest after study completion.

## Abbreviations

ABI: an inventory of coping with anxiety
ANOVA: Analysis of Variance
ANPS: Affective Neuroscience Personality Scales
ASI-3: Anxiety Sensitivity Index
BAS: Behavioral Approach System
BAT: Behavioral Avoidance Test
BIS: Behavioral Inhibition System
CERQ: the Cognitive Emotion Regulation Questionnaire
dB: decibel
ECG: electrocardiography
EMG: electromyography
FEAS: Questionnaire on Disgust and Fear of Spiders
FEE: Disgust Responsiveness
GAPED: Geneva Affective Picture Database
HC: Healthy control participants
Hz: hertz
ITI: Inter-Trial-Interval
m: meters
MINI: Mini International Neuropsychiatric Interview
ms: milliseconds
MWT-B: Multiple-Choice Vocabulary Intelligence Test
n: number of participants
NISS: Need Inventory of Sensation Seeking
NEO-FFI: NEO Five-Factor Inventory
PANAS: Positive and Negative Affect Schedule
RSES: Rosenberg Self-Esteem Scale
s: seconds
SCID-I: Structured clinical interview for the Diagnostic and Statistical Manual of Mental Disorders-IV
SCL: Skin Conductance Level
SEE: Scales for experiencing emotions
SP: Patients with spider phobia
SPQ: Spider Phobia Questionnaire
STAI: State-Trait-Anxiety-Inventory
UI-18: Intolerance of Uncertainty Scale

## Declarations

### Trial status

The trial (protocol version 1) was registered with Current Controlled Trials (ID: ISRCTN12918583) on 28^th^ February 2022. The recruitment process began at the 1^st^ of March 2022 and will finish on the 31^st^ of December 2023.

## Acknowledgements

The study is funded by the DGPs (German Society for Psychology: Biological Psychology and Neuropsychology) and Movisens (Peer-Mentoring-Program, awarded to FB and KR), and the Innovative Medizinische Forschung (IMF) of the medical faculty of the University of Münster (awarded to EJL, grant numbers: ME121805; LE121904). AH is supported by the German Research Foundation (Research Training Group 2493/1: 398510439).

Study materials were provided by Iris-Tatjana Kolassa and colleges and the Geneva Affective Picture Database (GAPED).

The study is sponsored by the University of Siegen (Adolf-Reichwein-Straße 2, 57072 Siegen, Germany; Contact: +49 271 740-4106).

The funders of the study are not involved in the collection, analysis, and interpretation of data. The sponsor of the study is involved in designing the study, and analysis and interpretation of data.

## Authors’ contributions

ASH, FB, EJL, JBF, LB, TK, UD, and KR designed the study. ASH, FB, and KR drafted the manuscript. ASH and FB are responsible for data collection and monitoring. All authors read and approved the final manuscript. The trial is carried out by the involved researchers and research assistants. While the involved researchers meet once every other month to check on data collection and management and on demand, all researchers responsible for data collection meet once a week to report on data collection and possible adverse events.

In case of important protocol modifications, all relevant parties (e.g., the respective researchers and trial registries) will be informed.

## Availability of data and materials

The responsible researchers and research assistants will have access to the final dataset. Data and statistical code will be provided upon reasonable request.

## Ethics approval and consent to participate

The study protocol was approved by the local Ethics Committee of the University of Siegen (07/2021), Germany (reference number: ER_39_2021). The study was designed in accordance with the Declaration of Helsinki, Good Clinical Practice guidelines, and the SPIRIT reporting guidelines. During the run of the study, ethical, legal, and social aspects are anticipated and addressed. Participants provide written informed consent to the experimental procedure before inclusion in the study. Informed consent is obtained by research assistants conducting the interviews for eligibility. Participation is entirely voluntary, and participants have the right to withdraw their consent at any time. Participant information is pseudonymized and stored on a secure database and can only be accessed by the involved researchers and research assistants.

## Randomization and Blinding

The randomization scheme was implemented utilizing block randomization. Participants are randomized via sequential numbers provided in a separate document.

Though participants are not especially informed about their allocation to a certain condition, they might have been aware of the condition. A blinding of investigators is not possible.

## Consent for publication

Not applicable.

The study protocol or parts of it have not been published elsewhere.

